# North American Pharmacist Licensure Examination Absolute Failures Warrant Monitoring of Colleges and Schools of Pharmacy

**DOI:** 10.1101/2024.04.09.24305491

**Authors:** John M. Pezzuto, Natalia Shcherbakova, Kimberly A. Pesaturo

## Abstract

Over the past three years, 7,978 graduates of pharmacy programs have failed the North American Pharmacist Licensure Examination (NAPLEX) on the first attempt. At present, the ACPE monitors programs with a passage rate of ≥2 standard deviations (SD) below the national mean pass rate. In 2023, this would lead to monitoring seven programs that produced 140 failures out of the total of 2,472 failures (i.e., 5.7%). In our view, this is neither equitable nor demonstrative of sufficient accountability. Analysis of failure counts among the 144 programs reported by NABP demonstrates a distribution curve highly skewed to the right. After evaluating average failure counts across all the programs, we suggest that schools with absolute failures ≥2 SD higher than the average number of failures of all programs should be flagged for monitoring. Based on the 2023 data, this corresponds to ≥35 failures/program. This threshold would flag 18 programs and 36.5% of the total failures. Of the seven programs that will be monitored based on the current Accreditation Council for Pharmacy Education criteria, only one would be captured by the ≥35 failure method of selection with the remaining six contributing only 85 total failures to the pool. Thus, if both criteria were to be applied, i.e., ≥35 failures and of ≥2 standard deviations below the national mean pass rate, a total of 24 programs would be monitored (16.6% of the 144 programs), that contribute 987 (39.9%) of the total failures.

## 1. Background

To gain licensure for practice following completion of the curriculum provided by an accredited health profession program, graduates must pass a board examination. Graduates from colleges and schools of pharmacy aspiring to enter the profession of pharmacy are expected to pass the North American Pharmacist Licensure Examination (NAPLEX^®^).^1^ It is reasonable to suggest that the overall NAPLEX performance of a graduating class reflects the ability of a professional degree program to supply student pharmacists adequately prepared for practice.^2^ Further, the first attempt passage rate should be considered most meaningful in terms of gauging the actual training provided by a respective degree program.^3^

Several reasons exist to desire high licensure exam passage rates. First, standardized tests are considered predictive of future performance.^4^ Higher NAPLEX scores are associated with higher pre-pharmacy and pre-advanced pharmacy practice experience grade point averages and on-time graduation rates.^5, 6^ Next, passage rate disclosure is mandated by accreditors from pharmacy and other professional programs.^7-9^ The current criterion applied by the Accreditation Council for Pharmacy Education (ACPE) for monitoring programs based on poor NAPLEX pass rate is to flag programs yielding a pass rate of ≥ 2 standard deviations below the national mean pass rate.^7^ This approach is more lenient than some other professional program standards; for example, the Accreditation Commission for Education in Nursing reviews examination pass rates for nursing professionals and requires either a minimum of 80% or greater for test-takers, or for mean pass rates to be at or greater than national or territorial means.^8^

## 2. Discussion

For numerous reasons, colleges and schools of pharmacy should be held accountable for the NAPLEX passage rates demonstrated by their graduates. The most recent data released by the National Association of Boards of Pharmacy (NABP), dated January 26, 2024, summarizes passing rates for 2021-2023 graduates.^10^ For 2023, the mean first time pass rate was 77.5% (SD 10.7%), and two standard deviations below the mean first time pass rate was 56.1%. Data are provided for 144 programs and 11,537 first-time test takers of which 2,472 graduates failed. According to the current ACPE policy, a program producing graduates with a pass rate that exceeds 56.1% would not be subjected to monitoring.^7^ Thus, if we include one program that achieved a pass rate of 56.3% (slightly above 56.1%), seven of the 144 programs will be monitored. Notably, these seven programs produced 140 failures out of the total of 2,472 failures (i.e., 5.7%).

In our view, there is an inequity in terms of this rationale for monitoring, and criteria beyond a pass rate less than or equal to two standard deviations below the national mean should be established to enhance program accountability. As an example, following the current monitoring paradigm, one program with a 50% pass rate contributing two failures will be monitored, and another program with a pass rate of 67.2% contributing 66 failures will not be monitored. This number of failures attributed to the latter program exceeds the number of graduates produced by over 50 other pharmacy programs, each with fewer than 66 total graduates. This is not a simple exercise in mathematics. It signals a problem.

The concern is not new. With NAPLEX pass rate data from 2015-2018, we proposed some alternative models by which pharmacy programs with poor passage rates should be considered for monitoring by the accrediting body.^11^ In essence, in addition to applying the current criterion of monitoring programs with passage rates ≥2 SD below the national mean, we proposed a maximum number of failures as a monitoring criterion. The maximum number of failures was not specifically prescribed, but it seemed most logical that it would fall in the range of 25 to >40 failures attributed to a single program.

Now, utilizing a report dated January 26, 2024, that summarizes passing rates for 2021-2023 graduates, ^10^ we thought it would be of interest to apply our strategy of identifying programs that should be considered for monitoring based on the concept of a maximum number of allowable failures for an individual program. Using the absolute number of failures emanating from first attempt test takers, Table 1 summarizes the number of programs meeting or exceeding the stated maximums.

**Table 1.** Comparison of ACPE school flagging rule with natural failure count approach.

| Schools flagged in 2023<br>based on ACPE Mean<br>minus 2 SD pass rate | Number of<br>schools (% total)<br>flagged if $n \geq 40$<br>failures | Number of<br>schools (%<br>total) flagged<br>if $n \geq 35$<br>failures | Number of schools<br>(% total) flagged if $n$<br>$\geq 30$<br>failures | Number of schools (%<br>total) flagged if $n \geq 25$<br>failures | Number of schools (% total)<br>flagged if $n \geq 20$ failures |
| --- | --- | --- | --- | --- | --- |
| $N = 7$ | $N = 6$ | $N = 12$ | $N = 18$ | $N = 25$ | $N = 43$ |
| Total $n$ failures = 140<br>(5.6%) | Total $n$ failures =<br>338 (13.7%) | Total $n$<br>failures = 564<br>(22.8%) | Total $n$ failures =<br>751 (30.4%) | Total $n$ failures = 942<br>(38.1%) | Total $n$ failures = 1334 (54.0%) |

We have now taken a closer look at the most recent data. The average failure rate among all 144 programs is 17.1 ± 11.8 (SD) failures/program. As shown in Figure 1,^12^ the data presents a right skewed distribution. Schools with failures of 56, 60, 62 and 66 students correspond to *z*-values of 3.30, 3.64, 3.81and 4.15 were removed as outliers. On the other hand, failure rates of 49 and 46 yield *z*-values of 2.71 and 2.45, respectively, and were left in place .^13^ Thus, after removing the four extreme values and evaluating the data from remaining 140 programs the mean (SD) now equals 15.8(9.3) failures per program.

**Figure 1.**
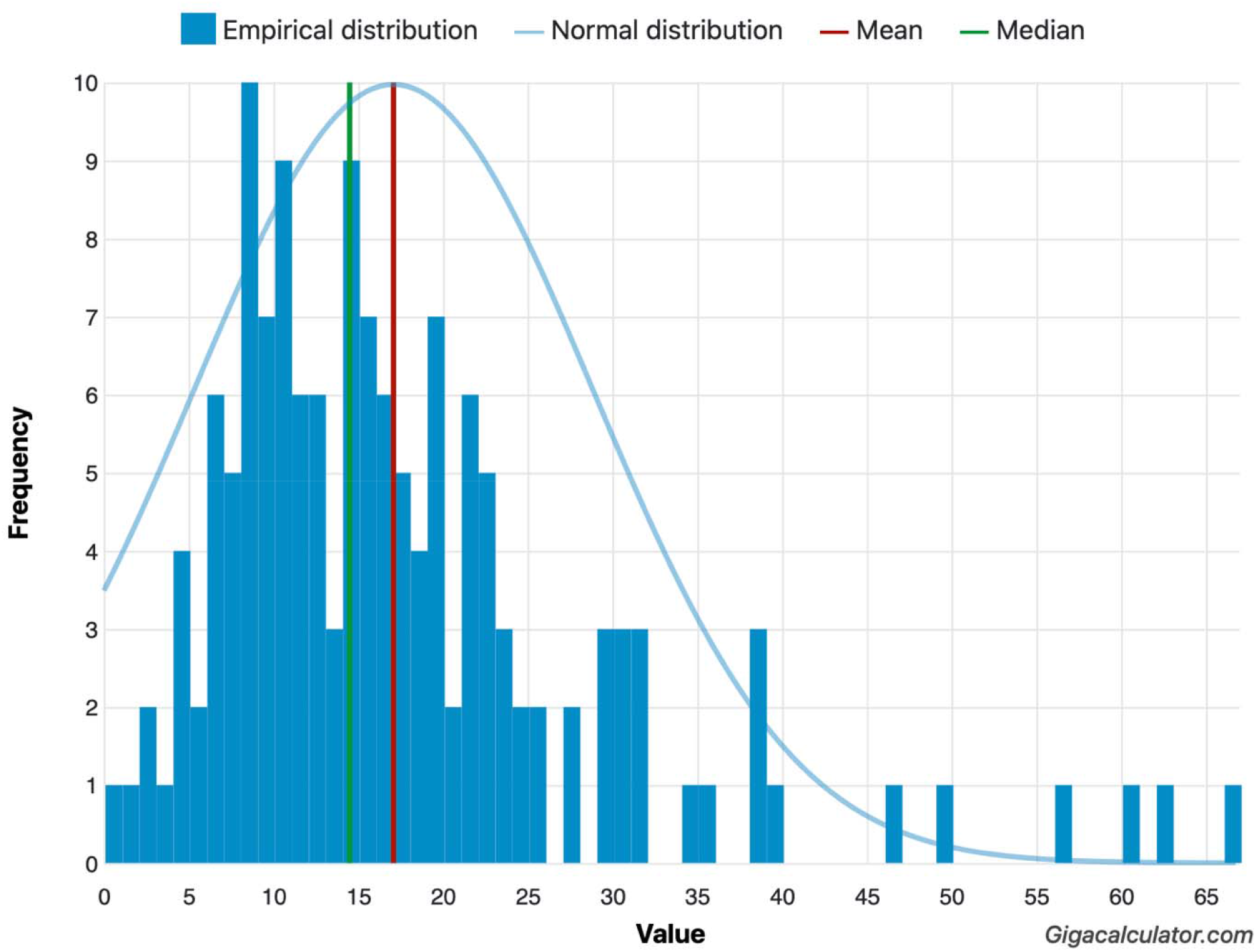
Distribution of failures attributed to each of the 144 colleges and schools of pharmacy.^1^ ^1^ Normality Calculator (Available at: https://www.gigacalculator.com/calculators/normality-testcalculator.php) was used to generate the chart. Value= absolute count of failures per school.

Applying the two standard deviation boundary as a criterion for monitoring a program, these data lead us to suggest that programs producing failures ≥ 2 SD higher than the average number of failures of all programs should be flagged for monitoring. Based on the 2023 data, this corresponds to ≥ 34.4 failures, which are captured in Table 1 under the heading of *n* ≥ 35 failures.

Accordingly, this threshold would flag 18 programs (≥ 35 and ≥ 40 failures) or 36.5% of all failures. Of the seven programs that will be monitored based on the current ACPE criterion, only one would be captured by the ≥ 35 failure method of selection. The remaining six programs currently flagged by ACPE contributed 85 failures to the pool. Thus, if both criteria were to be applied, i.e., ≥35 failures and of ≥2 standard deviations below the national mean pass rate, a total of 24 programs would be monitored (16.6% of the 144), that contribute 987 (39.9%) of the total failures.

It may be suggested that this approach is too stringent, or that the 2023 data may not be representative. However, consider the six programs that each produced over 40 graduates who failed. The average (SD) number of failures produced by these six programs for 2021, 2022 and 2023 were 57 (8), 72 (19), and 61 (14), respectively. Relative to 2021, there was no significant improvement in 2023 (*p* = 0.55), nor was there significant improvement relative to the 2022 failures (*p* = 0.11). This trend does not bode well for future improvement.

## 3. Conclusion

We feel this is a significant issue that needs to be addressed in the immediate future. It is obvious that institutions of higher education are under intense scrutiny by the public. There is a great deal of skepticism regarding the value of higher education in terms of return on investment, especially when finances and affordability are key factors in how adults perceive postsecondary education value.^14^ American Association of Colleges of Pharmacy surveys have indicated the student pharmacists incur an average debt exceeding $170,000 by the time of graduation.^15^ We realize a professional education is a partnership between the student and the institution. We realize not all graduates will pass NAPLEX on the first attempt. Nonetheless, the institution is the senior member of this partnership and needs to be held accountable within reasonable boundaries. It seems to us it is time to revisit and redefine the boundaries that are set based on NAPLEX pass rates.

## Data Availability

All data used are available online at https://nabp.pharmacy/wp-content/uploads/NAPLEX-Pass-Rates-2023.pdf

## Author Contributions

*Conceptualization*: All authors. *Writing–original draft*: All authors. *Writing–review & editing*: All authors.

## References

1. National Association of Boards of Pharmacy. The North American Pharmacist Licensure Examination. https://nabp.pharmacy/programs/examinations/naplex/. Accessed February 20, 2024.

2. National Association of Boards of Pharmacy. NAPLEX Competency Statements. https://nabp.pharmacy/programs/examinations/naplex/competency-statements/. Accessed February 20, 2024.

3. 2024 Candidate Application Bulletinx. North American Pharmacist Licensure Examination. https://read.nxtbook.com/nabp/bulletin/naplex_mpje_bulletin/what_is_the_naplex.html. Accessed February 20, 2024.

4. Kuncel NR, Hezlett SA. Standardized tests predict graduate students’ success. Science. 2007;315(5815):1080–1081.

5. Chisholm-Burns MA, Spivey CA, Byrd DC, McDonough SLK, Phelps SJ. Examining the association between the NAPLEX, Pre-NAPLEX, and pre- and post-admission factors. Am J Pharm Educ. 2017;81(5):article 86.

6. Maerten-Rivera J, Park SK, Fiano KS, Pavuluri N, Phillips J, Lebovitz L, et al. Multi-institutional analysis of student and program variables as predictors of performance on NAPLEX. Am J Pharm Educ 2022;86(4):article 8635.

7. Policies and procedures for ACPE accreditation of progressional degree programs. Accreditation Council for Pharmacy Education. https://www.acpeaccredit.org/pdf/CSPoliciesandProceduresJanuary2022.pdf. Accessed February 20, 2024.

8. 2023 accreditation manual. Accreditation Commission for Education in Nursing. https://resources.acenursing.org/space/AM/1842642947/2023+Accreditation+Manual. Accessed February 20, 2024.

9. ACOTE standards and interpretive guide. Accreditation Council for Occupational Therapy Education. https://acoteonline.org/accreditation-explained/standards/. Accessed February 20, 2024.

10. North American Pharmacist Licensure Examination® passing rates for 2021-2023 graduates. National Association of Boards of Pharmacy. https://nabp.pharmacy/wp-content/uploads/NAPLEX-Pass-Rates-2023.pdf. Accessed Feburary 20, 2024.

11. Shcherbakova N, Pezzuto JM. Monitoring of schools and colleges of pharmacy based on NAPLEX passage rate. Curr Pharm Teach Learn. 2023;15(10):854–860. 10.1016/j.cptl.2023.07.024.

12. Georgiev G.Z., “Normality Calculator”, [online] Available at: https://www.gigacalculator.com/calculators/normality-test-calculator.php Accessed Feb 14 2024.

13. Tabachnick BG, Fidell LS. Using multivariate statistics. 6th ed. Boston (MA): Allyn & Bacon/Pearson Education; 2013.

14. Munip L, Klein-Collins R. How they pay: the voices of adult learners on college affordability, and how institutions are respoinding. A CAEL research report. Council for Adult and Experiential Learning. https://files.eric.ed.gov/fulltext/ED629542.pdf. Accessed February 20, 2024.

15. American Association of Colleges of Pharmacy graduating student survey: 2021 national summary report. https://www.aacp.org/sites/default/files/2021-07/2021-gss-national-summary-report.pdf. Last accessed February 21, 2024.

